# Prospective development and validation of a liquid immune profile-based signature (LIPS) to predict response of metastatic cancer patients to immune checkpoint inhibitors

**DOI:** 10.1101/2020.08.03.20167163

**Authors:** Jian-Guo Zhou, Anna-Jasmina Donaubauer, Benjamin Frey, Ina Becker, Sandra Rutzner, Markus Eckstein, Roger Sun, Hu Ma, Philipp Schubert, Claudia Schweizer, Rainer Fietkau, Eric Deutsch, Udo S. Gaipl, Markus Hecht

## Abstract

**Background:** The predictive power of several novel biological markers for treatment response to immune checkpoint inhibitors (ICI) is still not satisfactory for the majority of patients with cancer. It should be the goal to identify easy available blood markers to predict early treatment response to ICI for the clinically highly relevant large number of metastatic cancer patients. The current interim analysis of patients of the ST-ICI cohort therefore focuses on the development and validation of a liquid immune profile-based signature to predict response of metastatic cancer patients to ICI targeting the programmed cell death protein 1 (PD-1)/PD-L1 axis.

**Methods:** A total number of 104 patients were prospectively enrolled. 54 immune cell subsets were prospectively analyzed in 89 patients’ peripheral blood by multicolor flow cytometry before the second administration of the ICI. Patients were randomly allocated to a training (n=56) and a validation cohort (n=33). Univariate Cox proportional hazards regression analysis and LASSO Cox model were used to create a predictive immune signature.

**Results:** 89/104 of the cancer patients provided whole blood samples. The identified liquid immune profile-based signature (LIPS) is based on five immune cell subtypes: CD14^high^ monocytes, CD8+/PD-1^+^ T cells, plasmacytoid dendritic cells (pDCs), neutrophils, and CD56^+^/CD16^+^ natural killer (NK)T cells. The signature found achieved a high accuracy (C-index 0.74 vs 0.71) for predicting overall survival (OS) benefit in both, the training and validation cohort. In both cohorts, the low-risk group had significantly longer OS than the high-risk group (HR 0.26, 95% CI: 0.12-0.56, p=0.00025; HR 0.30, 95% CI: 0.10 -0.91, p=0.024, respectively). Regarding the whole cohort, LIPS also predicted progression-free survival (PFS). Clinicopathological features with the exception of brain metastases didn’t affect the found LIPS, for both, OS and PFS.

**Conclusion:** Our study highlights the potential predictive value of LIPS based on easy available blood markers for immunotherapeutic benefit of metastatic cancer patients.

**Trial registration:** Prospectively registered in ClinicalTrials.gov (NCT03453892) on January 24, 2018.

## Introduction

Immune checkpoint inhibitors (ICI) can effectively restore the activity of exhausted CD8^+^ cytotoxic T cells and thereby trigger anti-tumor immune responses. During the last years, several ICI were approved for more than ten different tumor entities. However, still only the minority of patients show durable responses.^1^ In clinical trials, the expression of programmed cell death-ligand 1 (PD-L1) on tumor and/or immune cells has been frequently used to select patient subgroups with higher chances of treatment response. However, the predictive power of PD-L1 expression is restricted and its expression varies, as its expression is for example increased after radiotherapy. ^2-5^ Consequently, large effort has been made to identify routinely available blood and clinical markers that may predict response to ICI therapies,^6^ besides and complementary to analyses of genomic instability.^7^ The latter has been assessed via tumor mutational burden (TMB) ^8^ or mismatch repair deficiency.^9^ All of these methods have the potential to predict treatment responses to a certain extent, but are still complex to be used in the clinical routine. A further challenge is imaging of the cancer treatment response to ICI. Tumor lesions can initially increase in size due to inflammatory events.^10^ This pseudoprogression cannot easily be distinguished from an increase in size due to tumor growth. Consequently, the classical RECIST 1.1 criterial were modified to the iRECIST criteria, which recommend treatment beyond the first progression.^11^ The main problem of this procedure is that patients with real tumor progression loose time for a probably more efficient radio- and/or chemotherapy.

A recent strategy to complement these approaches is to analyze changes of peripheral blood immune cells at early time points during treatment.^12^ The peripheral blood immune status may have a high predictive power for treatment response of solid tumors to ICI, as several immune cells that were already proven to predict treatment responses when being present in the tumor tissue also circulate through the peripheral blood ^13^. Furthermore, drawing of peripheral blood can be repeated easily and without an additional risk for the patients compared to repeated biopsies of the tumor tissue. Immunophenotyping of peripheral blood from patients with stage IV melanoma before and after treatment with ICI already identified pharmacodynamic changes in circulating CD8^+^ T cells with an exhausted-phenotype. Clinical failure of the patients was identified to be associated with an imbalance between T-cell reinvigoration and tumor burden.^14^ Another study identified a predictive character of proliferating (Ki-67^+^) PD1^+^CD8^+^ T cells for response to anti-PD-1 immunotherapy in solid tumors.^15^ Using high-dimensional single-cell mass cytometry, Krieg and colleagues found HLA-DR high expressing monocytes to be of predictive value for responses to anti-PD-1 therapy.^16^

The here introduced preplanned biomarker analysis of the prospective ST-ICI cohort, the peripheral blood immune phenotype of metastasized cancer patients treated with ICI was prospectively monitored by multi-color flow cytometry. Our aim was to construct a predictive immune signature, based on changes of the peripheral blood immunophenotype that identifies at an early time point of therapy patients who benefit from ICI.

## Materials and Methods

### Patients

Metastatic cancer patients with clinically indicated treatment with ICI were eligible for this prospective non-interventional study. Patients could be included independent from cancer entity and concomitant radiotherapy. Criteria for eligibility were adult age of at least 18 years and the willingness of the patients to allow regular blood draws for immune phenotyping of peripheral blood. As the trial should represent the predominant clinical situation, there were no limitations regarding baseline Eastern Cooperative Oncology Group (ECOG) performance status or baseline routine blood parameters. Exclusion criteria were fertile patients who refused effective contraception during study treatment, persistent drug and/or alcohol abuse, patients not speaking German, patients in legal care and imprisoned patients.

### Study design and treatments

Patients that were treated with ICI directed against programmed cell death protein 1 (PD-1) or PD-L1 were selected from the ST-ICI cohort (NCT03453892). All ICI were indicated by the treating physician according to current guidelines and clinical standards. The following ICI were used: Nivolumab (Opdivo, Bristol-Myers Squibb, New York City, NY, USA), Pembrolizumab (Keytruda, Merck Sharp & Dohme, Kenilworth, NJ, USA), Atezolizumab (Tecentriq, Roche Co., Basel, Switzerland), Durvalumab (Imfinzi, AstraZeneca Group plc, Cambridge, England, U.K.), or Avelumab (Bavencio, Pfizer and Merck KGaA, Darmstadt, Germany). Dosing of the ICI was according to the European Medicines Agency (EMA) marketing authorizations.

The institutional review board at the Friedrich-Alexander-Universität Erlangen-Nürnberg approved the study (number: 2_17 B). The study was performed in accordance with the Declaration of Helsinki. All patients gave written informed consent before enrollment that comprised a data privacy clause for data collection and analysis for research purpose. The research design for the identification of a predictive signature for cancer patients treated with ICI is displayed in **Figure 1A**. The redaction of the manuscript followed the STROBE guidelines for observational studies.

**Figure 1.**
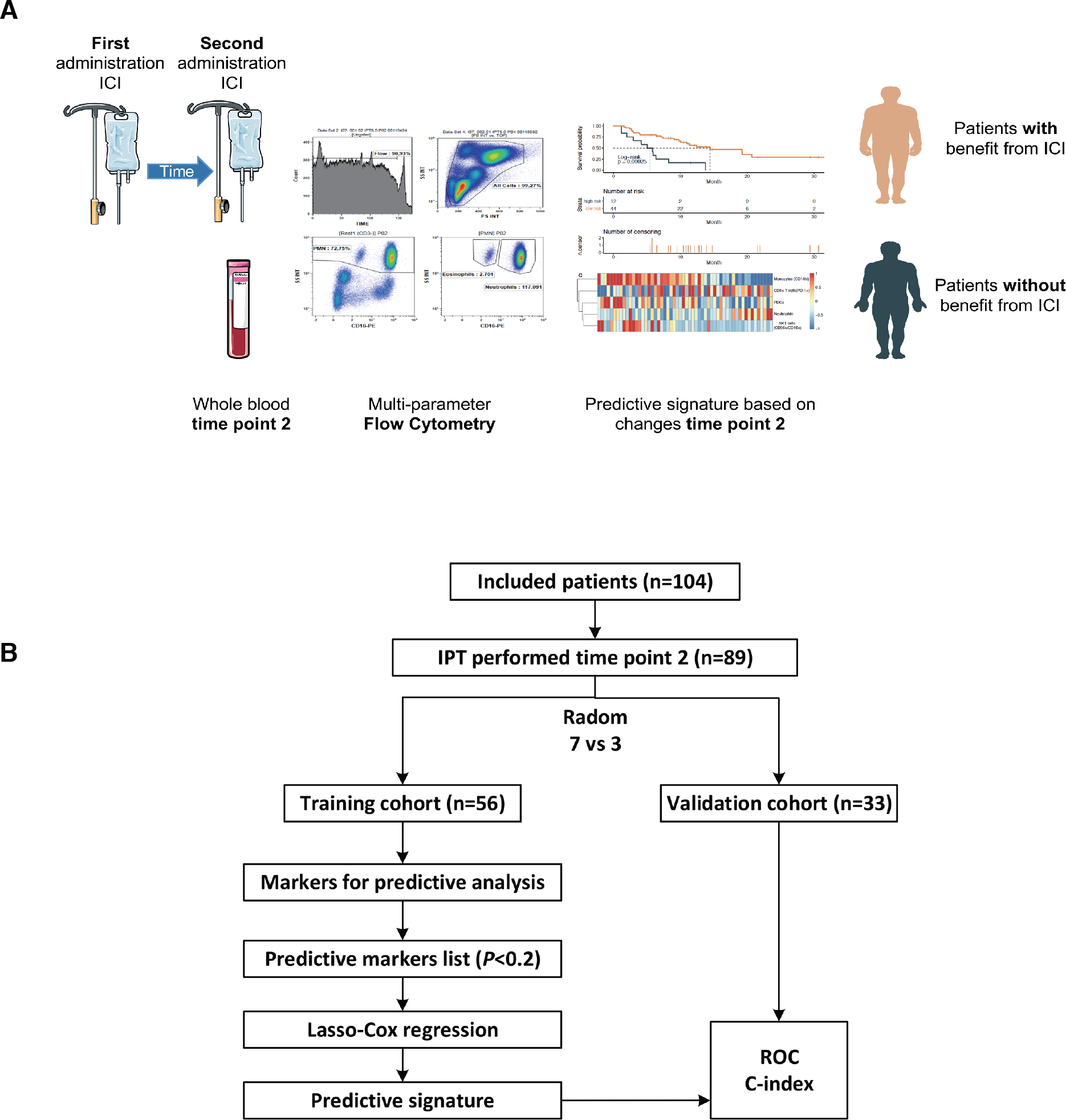
Research design (A) and flowchart (B) for the identification of a predictive signature by immunophenotyping (IPT) for metastatic cancer patients treated with immune checkpoint inhibitors (ICI).

### Endpoints and assessments

The primary endpoint of the here presented prospective interim analyses was the association between changes of the patients’ peripheral blood immunophenotypes and the clinical outcomes. Main clinical outcome parameters were overall survival (OS), progression-free survival (PFS). OS and PFS were calculated starting with the first administration of the ICI. Peripheral whole blood immune phenotyping was performed before each administration of the ICI. The current analysis focused on the early immune phenotypes, namely before the second administration of the ICI.

### Multi-colour flow cytometry

Whole blood samples of the patients were collected and the fresh whole blood was analyzed by multi-colour flow cytometry according to our previously published and optimized immunophenotyping (IPT) protocols.^17 18^ IPT was performed within three hours after the collection of whole blood. Data acquisition was performed on a Gallios Flow Cytometer (Beckman Coulter) in the standard filter configuration. The Kaluza® Flow Analysis Software (Beckman Coulter) was used for data analysis. The immune cell subpopulations analyzed are visualized and specified in **sFigure 1**.

### Data collection

Differences in OS and PFS were analysed in dependence of the absolute immune cell counts of each immune cell subset and in dependence of the clinical factors age, gender, tumour entity, PD-L1 expression, and the presence of brain metastases.

### Development and validation of liquid immune profile-based signature (LIPS)

The overall workflow for the development of the LIPS is shown in **Figure 1B**. Univariate Cox proportional hazard regression was applied to examine the association between peripheral blood immunophenotypes and patients’ OS.^19^ Peripheral blood immune cells statistically significant associated (P < 0.2) with OS of the patients served as candidates for further analyses. To uncover the practicability and accuracy of LIPS for metastasized cancer patients treated with ICI, all patients were divided randomly into the training (70%) and validation (30%) cohorts. Least absolute shrinkage and selection operator (LASSO) could reduce the complexity of the model and can well be used with the Cox proportional hazard regression model for survival analysis with high dimensional data.^20 21^ The Cox-LASSO regression model was applied to develop a multi-immunophenotype-based predictive signature for OS in the training cohort. After 200,000-time steps for LASSO, the best c-index related model was selected as LIPS. Subsequently, we analyzed the data in a validation cohort for this model, to assess its feasibility and reliability in patients treated with ICI. The optimal cut off value for the risk score (**Figure 2A and D**) was determined by *surv_cutpoint* function in *survminer* package. All patients were divided into different groups (high-risk or low-risk) based on the cut-off of the risk score, which was calculated by considering the expression of immunophenotypes and the correlation coefficient. In order to validate the predictive ability of LIPS for PFS, these same coefficients of LIPS were calculated as risk score for survival analysis.

**Figure 2.**
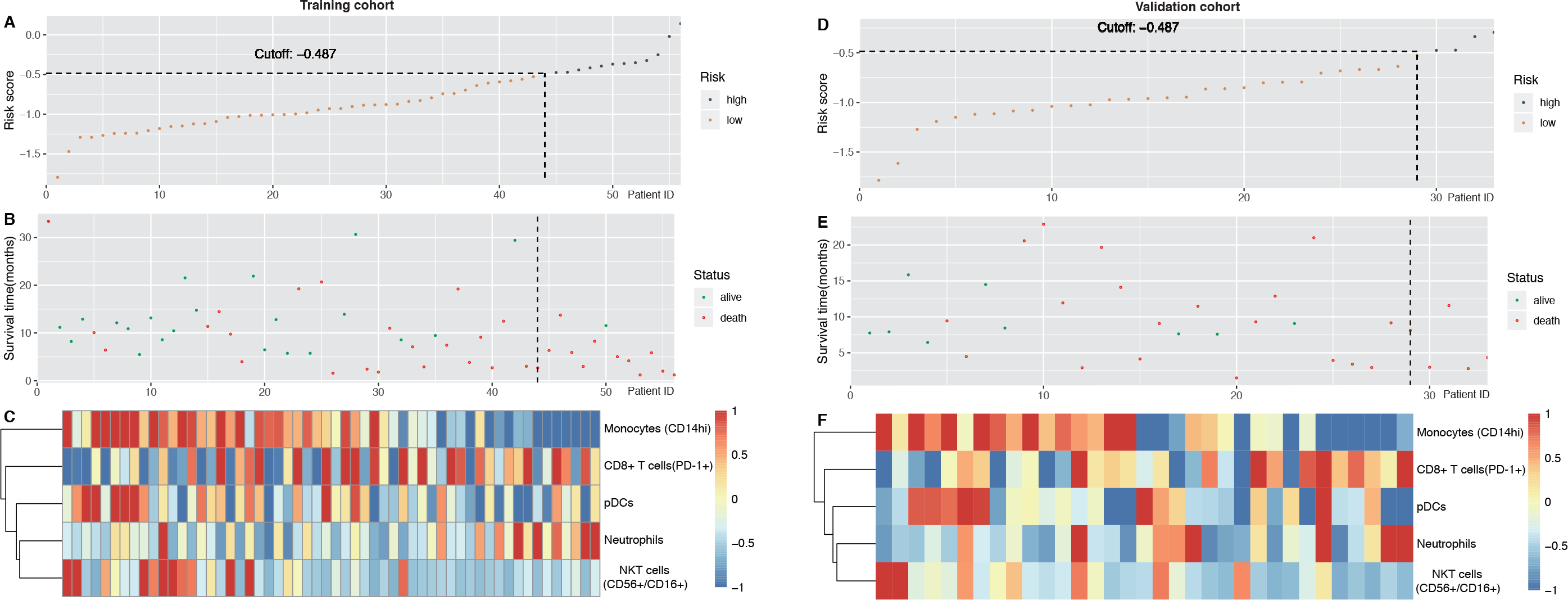
Characteristics of the liquid immune profile-based signature (LIPS) in the training and validation cohorts. **(A)** The risk score of each metastatic cancer patient (patient ID) treated with immune checkpoint inhibitors (ICI) in the training cohort. **(B)** Overall survival and survival status of metastatic cancer patients in the training cohort. **(C)** Heat map of immune cell counts of metastatic cancer patients in the training cohort. **(D)** The risk score of each metastatic cancer patient treated with ICI in the validation cohort. **(E)** Overall survival and survival status of metastatic cancer patients in the validation cohort. **(F)** Heat map of immune cell counts of metastatic cancer patients in the validation cohort. pDCs: plasmacytoid dendritic cells; NKT cells: natural killer T cells; PD-1: Programmed cell death protein 1.

### Statistical analysis

Associations between clinical characteristics in the training and the validation cohorts were evaluated using the chi-square test. OS time was defined from the date of the first administration of the ICI to the date of the last follow-up or death of the patient. PFS time was defined from the date of the first administration of the ICI to the date of the last follow-up or first radiological confirmed progression (e.g. imaging date) or date of death (whichever occurs first). The Kaplan-Meier method and Cox proportional hazard regression models were applied to compare survival of the different groups with the immunophenotypes, LIPS and related clinical factors. Univariate, multivariate and subgroup analyses were used to evaluate the impact of other confounding factors. Results of Cox regression analysis are described by means of hazard ratios (HR), 95%CI of HR and *P* values (Wald test). The concordance index (C-index) and the time-dependent receiver operating characteristic (ROC) curve, and the area under the ROC curve (AUC) values were calculated for different models as a measure of the discriminatory ability that allows comparison of signatures. A signature with a C-index of 0.5 has no predictive value, but the signature with a C-index of 1 would allow a perfect prediction of the patient’s outcome.^22^ The C-index was analyzed using the *survcomp* (version 1.22.0).^23^ The ROC curve and AUC values were calculated with the *timeROC* package (version 0.3); for survival analyses the *survival* package was used. All of the analyses were carried out using R version 3.6.1 (R Foundation for Statistical Computing) and related packages. *P* ≤ 0.05 was considered to be statistically significant.

## Results

### Patient characteristics

A total of 104 patients were prospectively enrolled in the ST-ICI cohort between April 2017 and August 2019 (**Figure 1B**). Whole blood samples for immunophenotyping before the second ICI administration were available of 89 patients. The patient characteristics are presented in **Table 1**. The median age was 65.8 years; 73% were male. Most frequent tumor entities were head and neck squamous cell carcinoma (HNSCC) in 40 patients (45%) and non-small cell lung cancer (NSCLC) in 39 patients (44%). The used drugs were Nivolumab in 58 patients (65%), Pembrolizumab in 20 patients (23%), Durvalumab in 8 patients (9%), Avelumab in 2 patients (2%), Atezolizumab in 1 patient (1%).

**Table 1.** Baseline characteristics of metastatic cancer patients treated with ICI and for which immunophenotyping was performed before the second administration of ICI.

| Features | Training cohort | Validation cohort | P-value |
| --- | --- | --- | --- |
| **Total** | 56 | 33 |  |
| **Gender** |  |  | 0.5571 |
| Male | 42 (75%) | 23 (69.7%) |  |
| Female | 14(25%) | 10(30.3%) |  |
| **Age** |  |  | 0.4133 |
| >=60 | 16(28.57%) | 13(39.39%) |  |
| <60 | 40(71.43%) | 20(60.61%) |  |
| **Tumor entity** |  |  | 0.8186 |
| HNSCC | 24(42.86%) | 16(48.49%) |  |
| NSCLC | 26(46.42%) | 13(39.39%) |  |
| others | 6(10.71%) | 4(12.12%) |  |
| **Brain metastasis** |  |  | 0.6538 |
| Yes | 12(21.43%) | 5(15.15%) |  |
| No | 44(78.57%) | 28(84.85%) |  |
| **PD-L1 expression** |  |  | 0.8995 |
| <1% | 19(33.93%) | 12(36.37%) |  |
| 1-49% | 18(32.14%) | 10(30.3%) |  |
| 50%-100% | 17(30.36%) | 11(33.33%) |  |
| NA | 2(3.57%) | 0 |  |

The ICI was first line treatment in 19 patients (21%). In the recurrent and/or metastatic setting, 13 patients (15%) had received one prior systemic treatment and 57 patients (64%) two or more prior systemic treatments. The median follow up was 8.3 months. A total of 57 OS-events and 72 PFS-events occurred during the follow up period. Median OS was 8.7 months, median PFS was 4.2 months.

### Development and definition of the LIPS

After the data cleaning steps, 54 peripheral blood immune markers were included into an univariate cox survival analysis. The following 14 of them were associated with OS (*P*< 0.2): monocytes (CD14^high^), monocytes (CD14^low^), neutrophils, dendritic cells (DCs), myeloid (m)DCs-1, mDCs-2, plasmacytoid DCs (pDCs), natural killer (NK) cells (CD56^high^/CD16^+^), NKT cells (CD56^+^/CD16^+^), NKT cells (CD16^+^), CD8+ T cells (PD-1^+^), CD8^+^ T cells (CD25^+^), CD8^+^ T cells (CD69^+^), and regulatory T cells (Tregs) (**Table 2**).

**Table 2.** Univariate Cox regression (p<0.2).

| Characteristics | HR | 95% CI | P Value |
| --- | --- | --- | --- |
| **Monocytes (CD14<sup>high</sup>)** | 0.28 | 0.16 - 0.47 | 0 |
| **Monocytes (CD14<sup>low</sup>)** | 1.37 | 1.14 - 1.66 | 0.00109 |
| **Neutrophils** | 1.68 | 1.15 - 2.44 | 0.00682 |
| **DCs** | 0.69 | 0.47 - 1.03 | 0.06665 |
| **mDCs-1** | 0.47 | 0.27 - 0.83 | 0.00991 |
| **mDCs-2** | 0.75 | 0.58 - 0.97 | 0.0258 |
| **pDCs** | 0.56 | 0.4 - 0.8 | 0.00114 |
| **NK cells (CD56<sup>high</sup>/CD16<sup>+</sup>)** | 0.84 | 0.64 - 1.09 | 0.19433 |
| **NKT cells (CD56<sup>+</sup>/CD16<sup>+</sup>)** | 0.61 | 0.4 - 0.92 | 0.01906 |
| **NKT cells (CD16<sup>+</sup>)** | 0.76 | 0.58 - 1 | 0.0491 |
| **CD8+ T cells (PD-1<sup>+</sup>)** | 1.63 | 1.06 - 2.51 | 0.02604 |
| **CD8+ T cells (CD25<sup>+</sup>)** | 1.2 | 0.93 - 1.56 | 0.1591 |
| **CD8+ T cells (CD69<sup>+</sup>)** | 0.75 | 0.49 - 1.15 | 0.18642 |
| **T<sub>regs</sub>** | 0.79 | 0.58 - 1.08 | 0.14495 |

The ST-ICI cohort was then randomly assigned to a training cohort (n=56) and a validation cohort (n=33) for further LASSO proportional hazards Cox regression analysis. After 200,000 times repeat LASSO regression, the highest C-index’s model was chosen as the LIPS. This predictive signature model obtained from the training cohort showed the following risk score = (0.00001219963×the absolute immune cell counts of neutrophils) + (−0.0335468×the absolute immune cell counts of pDCs) + (−0.08766993 × the absolute immune cell counts of NKT cells (CD56^+^/CD16^+^)) +(−0.01120408 × the absolute immune cell counts of monocytes (CD14^high^)) + (0.004149556× the absolute immune cell counts of CD8^+^ T cells (PD-1^+^)). In this signature, neutrophils and CD8^+^ T cells being positive for PD-1 surface expression were positive coefficients, which means that patients with increasing absolute counts had a shorter OS. However, pDCs, NKT cells (CD56^+^/CD16^+^) and monocytes (CD14^high^) were negative coefficients, which means that patients with increasing absolute cell counts had a longer OS.

According to best C-index (0.74, 96%CI 0.67-0.82), the optimal cut-off is −0.487, which classified the training cohort into a low-risk group (n=44) and a high-risk group (n=12) (**Figure 2 A-C**).

In the training cohort, each unit increase in LIPS was associated with a 0.26 fold increase in OS rates (95%CI: 0.12-0.56, P =0.00025). Median OS in the LIPS low- and high-risk groups was 14.5 (95% CI: 10.1-NE (not estimable)) vs 5.4 (95% CI: 3.0-NE) months, respectively (**Figure 3A**).

**Figure 3.**
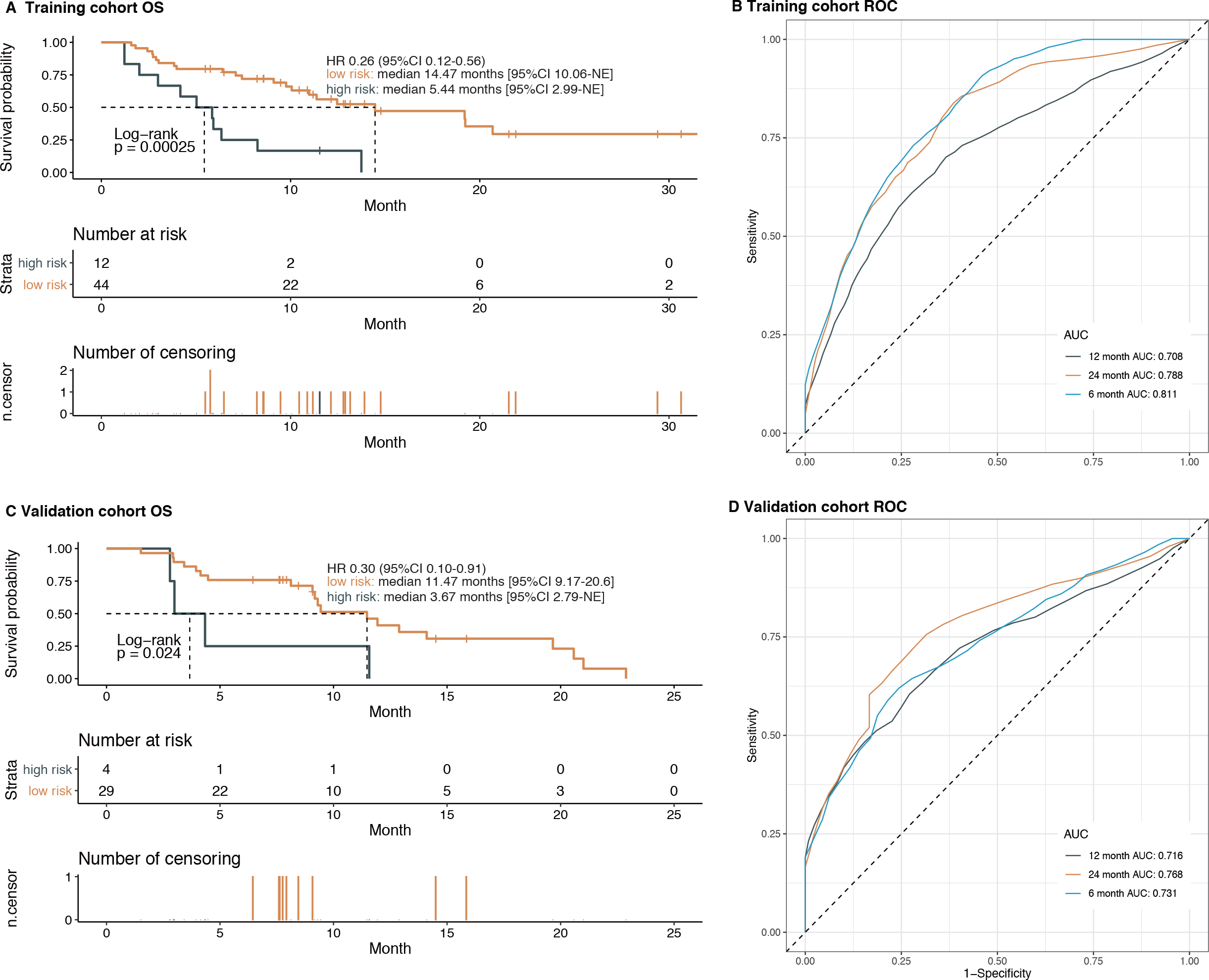
The ability of the LIPS to predict the overall survival in the training and validation cohorts. **(A)** The overall survival in training cohort stratified by the LIPS into high- and low-risk with the *P*-value. **(B)** Time-dependent ROC curves of LIPS in the training cohort. **(C)** The overall survival in validation cohort stratified by the LIPS into high- and low-risk with the *P*-value. **(D)** Time-dependent ROC curves of LIPS in the validation cohort.

The validation cohort was classified using the same cut-off of LIPS into a low-risk group (n=29) and a high-risk group (n=4) (**Figure 2D-F**). The validation cohort confirmed the proposed risk model (C-index=0.71, 95%CI: 0.62-0.81). The median OS in the low- and high-risk groups was 11.5 (95% CI: 9.2-20.6) vs 3.7 (95% CI: 2.8-NE) months (HR=0.30, 95% CI: 0.10–0.91, P = 0.024), respectively (**Figure 3C**).

The sensitivity and specificity of LIPS for predicting the OS were plotted in a time-dependent ROC. In the training cohort, the AUC values for 6-, 12- and 24-month OS prediction were 0.811, 0.708 and 0.788 respectively (**Figure 3B**). In the validation cohort, the AUC values for 6-, 12- and 24-month OS prediction were 0.731, 0.716 and 0.768, respectively (**Figure 3D**).

In the total ST-ICI cohort, median OS in the low- and high-risk groups was 12.5 (95% CI: 9.8-19.7) vs 4.7 (95% CI: 3.0-NE) months (HR=0.28, 95% CI: 0.15–0.52, P<0.001), respectively (**Figure 4A**). The time-ROC results indicate that this signature is still a powerful predictor for OS in the whole cohort (AUC_6m_=0.77, AUC_12m_=0.72, AUC_24m_=0.82) (**Figure 4B**).

**Figure 4.**
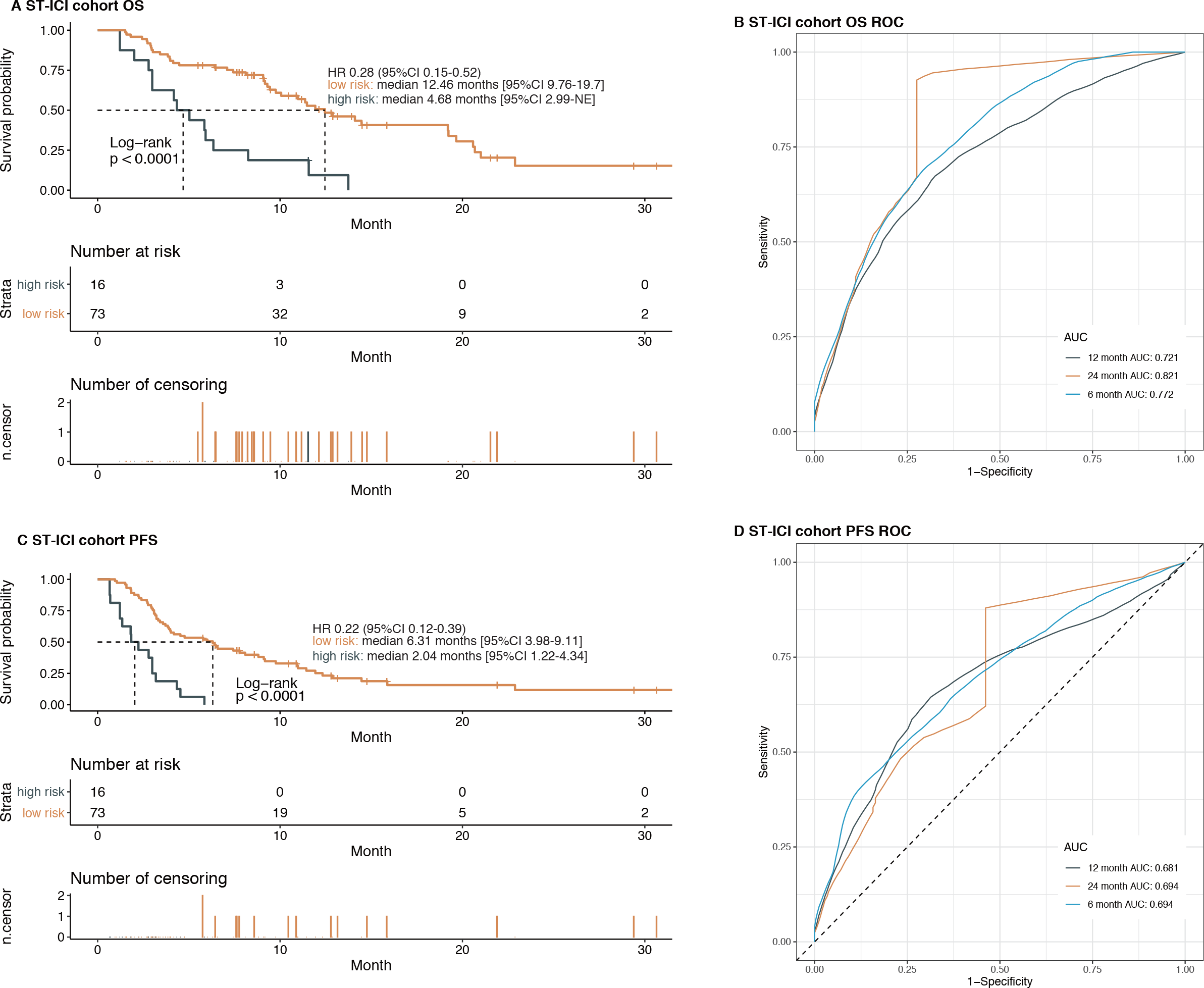
LIPS predicts survival benefit from ICI treatment in all metastatic cancer patients of the ST-ICI cohort. **(A)** The overall survival in all patients stratified by the LIPS into high- and low-risk with the *P*-value. **(B)** Time-dependent ROC curves of LIPS for overall survival in all patients. **(C)** The progression-free survival in all patients stratified by the LIPS into high- and low-risk with the *P*-value. **(D)** Time-dependent ROC curves of LIPS for progression-free survival in all patients.

Additionally, LIPS also can predict progression-free survival benefit for patients treated with ICI (low-risk vs high-risk: HR=0.22 [95%CI 0.12-0.39]; the median PFS of low risk and high risk are 6.3 and 2.0 months, respectively (**Figure 4C**); AUC_6m_=0.69, AUC_12m_=0.68, AUC_24m_=0.69 (**Figure 4D**). These results indicate that the developed LIPS can predict which cancer patients have prolonged OS and PFS after treatment with anti-PD-1 or anti-PD-L1 in the early phase.

### Predictive role of LIPS in metastatic cancer patients treated with ICI

To determine whether the LIPS could serve as an independent predictive factor, Cox proportional hazard regression model was used for the detection of the relationships between OS and the clinical factors (**Table 3**). In the ST-ICI cohort, univariate analysis showed that LIPS and brain metastasis were significantly associated with OS, while multivariate analysis showed that age, PD-L1 expression 50-100 % and LIPBS significantly associated with OS (P < 0.05).

**Table 3.** Univariate and multivariate Cox regression analysis for overall survival and progression-free survival.

| Parameter | Overall Survival |  |  |  |  |  | Progression-Free Survival |  |  |  |  |  |
| --- | --- | --- | --- | --- | --- | --- | --- | --- | --- | --- | --- | --- |
|  | univariate Cox regression |  |  | multivariate regression |  |  | univariate regression |  |  | multivariate regression |  |  |
|  | HR | 95%CI | P | HR | 95%CI | P | HR | 95%CI | P | HR | 95%CI | P |
| **LIPS signature** |  |  |  |  |  |  |  |  |  |  |  |  |
| High risk | Ref |  |  | Ref |  |  | Ref |  |  | Ref |  |  |
| Low risk | 0.28 | 0.15-0.52 | <0.001*** | 0.27 | 0.12-0.57 | <0.001*** | 0.22 | 0.12-0.39 | <0.001*** | 0.23 | 0.10-0.52 | <0.001*** |
| **Age** |  |  |  |  |  |  |  |  |  |  |  |  |
| < 60 yr | Ref |  |  | Ref |  |  | Ref |  |  | Ref |  |  |
| ≥ 60 yr | 1.36 | 0.77-2.41 | 0.3 | 2.22 | 1.12-4.41 | 0.023* | 1.13 | 0.68-1.86 | 0.6 | 1.56 | 0.89-2.75 | 0.12 |
| **Gender** |  |  |  |  |  |  |  |  |  |  |  |  |
| Male | Ref |  |  | Ref |  |  | Ref |  |  | Ref |  |  |
| Female | 1 | 0.54-1.86 | 1 | 1.46 | 0.68-3.09 | 0.323 | 0.77 | 0.44-1.34 | 0.4 | 0.98 | 0.52-1.86 | 0.95 |
| **Tumour entity** |  |  |  |  |  |  |  |  |  |  |  |  |
| Others | Ref |  |  | Ref |  |  | Ref |  |  | Ref |  |  |
| HNSCC | 0.91 | 0.54-1.55 | 0.7 | 0.54 | 0.23-1.31 | 0.172 | 1.41 | 0.88-2.25 | 0.1 | 0.81 | 0.34-1.88 | 0.618 |
| NSCLC | 0.94 | 0.56-1.59 | 0.8 | 0.6 | 0.25-1.46 | 0.263 | 0.7 | 0.44-1.12 | 0.1 | 0.56 | 0.23-1.38 | 0.208 |
| **Brain metastasis** |  |  |  |  |  |  |  |  |  |  |  |  |
| Yes | Ref |  |  | Ref |  |  | Ref |  |  | Ref |  |  |
| No | 0.43 | 0.24-0.79 | 0.005** | 0.52 | 0.25-1.1 | 0.087 | 0.46 | 0.26-0.80 | 0.005** | 0.75 | 0.34-1.66 | 0.481 |
| **PD-L1 expression** |  |  |  |  |  |  |  |  |  |  |  |  |
| < 1 % / NA | Ref |  |  | Ref |  |  | Ref |  |  | Ref |  |  |
| 1-49 % | 0.69 | 0.39-1.24 | 0.2 | 0.57 | 0.30-1.12 | 0.106 | 0.61 | 0.36-1.02 | 0.06 | 0.51 | 0.27-0.96 | 0.036* |
| 50-100 % | 0.78 | 0.43-1.41 | 0.4 | 0.44 | 0.20-0.93 | 0.031* | 0.89 | 0.54-1.47 | 0.6 | 0.61 | 0.31-1.18 | 0.142 |

**Note:** \* p<0.05, \*\* p<0.01, \*\*\*p<0.001

Furthermore, LIPS and PD-L1 expression 1-49 % were significantly associated with PFS in univariate and multivariate analysis. These results indicate that LIPS remained an independent survival benefit predictor for metastatic cancer patients treated with ICI.

### Applicability of LIPS in cancer patients with different clinical characteristics

Post-hoc subgroup analyses based on patient characteristics suggested that OS and PFS hazard ratios favoured the LIPS low risk group in most subgroups (**Figure 5, sFigures 2-6**). Regarding OS, the subgroup analyses by PD-L1 expression suggested that median OS tended to be longer in patients with PD-L1 expression < 1% in the low-risk group (HR= 0.45 (95%CI 0.16-1.24), 9.1 months [95% CI 4.5–21] vs 5.0 months [3.0–NE]; **Figure 5A, sFigure 3**). However, no differences of OS and PFS were found between low-risk and high-risk groups in patients with brain metastasis (**sFigure 4**). Median OS and PFS of patients stratified by tumour entity, age, and gender are shown in the **sFigures 2, 5**, and **6**, respectively.

**Figure 5.**
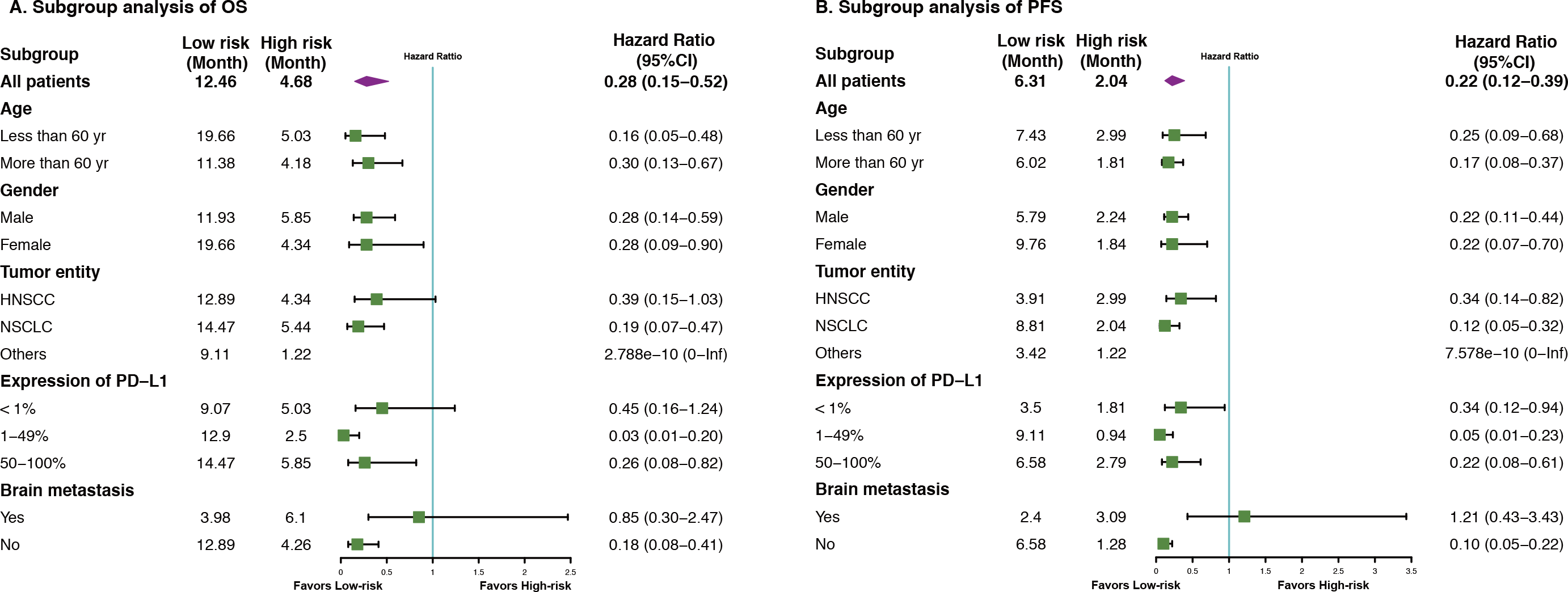
Subgroup analysis of the LIPS predict survival benefit of patients treated with ICI by baseline characteristics. Subgroup analysis of the LIPS predict overall survival benefit **(A)** and progression-free survival benefit **(B)** of patients treated with ICI. Hazard Ratio and 95% Confidence Intervals.

## Discussion

In recent years, ICI with or without chemotherapy were approved for multiple recurrent or metastatic cancer types. However, a maximum of 20% of non-selected patients experience long-term benefit.^2 24^ Numerous studies suggest that TMB may predict clinical response to ICI across multiple cancer types.^25-27^ However, different studies used different cut-off values based on independently companies’ database or cancer types to define low and high TMB.

The detection of PD-L1 by different methods or cut-off values could also guide anti-PD-1/PD-L1 therapy, but still harbours the risk of a false-negative patient stratification.^28-30^ The gene methylation status may also serve to select patients who will experience clinical benefit from PD-1 blockade.^31^ Nevertheless, there is still particular need of additional or complementary timesaving, cost-effective, safe and easy applicable tools to identify cancer patients who should benefit from ICI.

In this context, a detailed knowledge about the immune status, both in the tumor and in the periphery, is needed to judge about the potential of cancer immunotherapy and the efficacies of immunotherapies.^32^ Tumor immunogenicity scores are under evaluation as predictors for responses to ICI.^33^ Previous studies already indicated peripheral blood immune cell subsets like PD-1^+^CD56^+^ T cells ^34^ or CD4+ and CD8+ cells ^35^ to be associated with favourable outcome for advanced melanoma patients treated with ipilimumab directed against cytotoxic T lymphocyte antigen-4 (CTLA-4). Now we identified a liquid immune profile-based signature to early predict response of metastatic cancer patients to anti-PD-1/PD-L1 treatment.

To our best knowledge, the prospective ST-ICI cohort is the first one, which prospectively included a whole-blood multicolor flow cytometry-based approach for identification of a detailed peripheral immune status in a clinical relevant multi-type advanced cancer patient cohort treated with anti-PD-1/PD-L1 ICI. To avoid large volume blood sampling for preparation of peripheral blood mononuclear cells (PBMC) and to not exclude granulocytes from the analyses, the IPT was performed in fresh whole blood without preceding isolation of specific cells. Thus, whole blood assays gain importance in easy screening of large number of blood parameters. Within the ST-ICI cohort, 54 immune cell subsets were monitored in the patients with the need of less than 1 ml of peripheral whole blood. Our identified LIPS has the advantage of being easily integrated into clinical use for nominal expense. In a first step, 14 immune cell subsets were identified to be associated with OS. Furthermore, LASSO regression constructed the LIPS signature including neutrophils, pDCs, NKT cells (CD56^+^/CD16^+^), monocytes(CD14^high^) and CD8^+^T cells (PD-1^+^). This signature includes innate and adaptive immune cells and serves as an effective tool for the identification of metastatic cancer patients who benefit from ICI treatment. It is becoming more and more evident that multiple immune cell subsets in a concerted action may help for selecting patients that are likely to respond to ICI.^36^

We identified that increasing amounts of neutrophils in the peripheral blood of cancer patients were associated with less benefit from ICI treatment. Neutrophils have been reported to support for example the development of metastasis through multiple mechanisms, such as the release of proteases that degrade antitumor factors, and leukotrienes that propagate metastasis-initiating cells. In addition, pro-tumorigenic neutrophils suppress antitumor T cell responses.^37^ Decreased migration of neutrophils to tumor areas or the inhibition of granulocyte colony stimulating factor to decrease the amount of neutrophils has already shown efficacy in preclinical models.^38^ Recent findings suggests that neutrophil antagonism will improve the efficacy of ICI therapy in the future.^12^

Regarding innate immune cells not of the granulocytic, but of the monocytic compartment, increased amounts of CD14 high expressing monocytes in the peripheral blood are associated with improved prognosis. This is in accordance with the findings of Krieg et al who identified CD14^+^/HLA-DR^+^ monocytes as a strong predictor of PFS and OS in response to anti-PD-1 immunotherapy in melanoma patients.^39^ We here show the importance of CD14 high expressing monocytes in the peripheral blood for non-melanoma solid tumour patients.

Further, we found that increased amounts of plasmacytoid dendritic cells (pDCs) are beneficial for the PFS and OS. pDCs have a central role in activating host innate and adaptive immune responses and they are known as the major IFN type I-producing cells. Thereby, pDCs activate many other cell types, such as monocytes, NK cells, and T cells which are known to be also central for anti-tumor immune responses.^40^ High levels of circulating pDCs were already found to be predict a favorable outcome in patients with breast cancer ^41^ and for patients with pancreatic adenocarcinoma.^42^ NKT cells are suggested to induce a cross talk of pDCs with conventional DCs that result in induction of memory CD8+ T cells.^43^ We identified increased numbers of NKT cells in patients of the ST-ICI cohort to be favorable for PFS and OS. Just recently, it was shown for patients with NSCLC that increased amounts of peripheral NK cells correlate with responses to anti-PD-1 treatment.^44^ The role for NKT cells in this scenario has not been investigated before, but it is becoming more and more evident that NKT cells are important in anti-tumor immunity as they e.g. reinvigorate exhausted immune cells in the tumor microenvironment.^45^

Exhausted immune cells fail to contribute to anti-tumor immunity. Increased numbers of PD-L1^+^ NK cells in the peripheral blood, e.g., are associated with poor response to anti-PD-1 treatment of patients with NSCLC.^46^ However, intratumoral CD8^+^ T cells which are positive for PD-1^+^ were shown to be predictive for response and survival upon anti-PD-1 antibody therapy, as shown in a small cohort of patients with NSCLC.^47^ Studies in chronic myeloid leukemia patients just recently revealed that differences in PD-1 Expression on CD8^+^ T-cells might predict the disease course.^48^ Our immune monitoring suggests that increased amounts of circulating PD-1^+^ CD8^+^ T cells at early time points of ICI therapy predict a poorer OS and PFS in patients with metastatic solid tumors treated with ICI. This might indicate that in these cases the activated CD8^+^ T cells cannot enter properly into the tumor and exert their anti-tumor activity.

The ST-ICI cohort composes a clinically highly relevant group of advanced stage patients with solid tumors, what includes some confounding factors like treatment with different ICI, different tumor types, brain metastasis, and PD-L1 expression. However, the multivariable Cox regression and subgroup analyses suggests that LIPS is an independent prognostic factor for advanced cancer patients treated with ICI targeting the PD-1/PD-L1 axis. Subgroup analysis suggest that this immune signature can stably predict OS and PFS benefit for cancer patients treated with ICI in any subgroups, except for brain metastasis. Although patients with brain metastases can benefit from ICI treatment,^49^ the knowledge about the immune microenvironment at this tumor location side is scarce,^50 51^ and future trials will have to particularly focus on this subgroup of the ST-ICI study population. In present clinical practices, PD-L1 expression is an essential biomarker as drug approvals mostly depend on its expression. However, our subgroup analyses revealed that the LIPS predicts survival benefits independent from the PD-L1 expression status. Notably, LIPS identified more than 82% of all advanced cancer patients, who benefit from anti-PD-1/PD-L1 antibodies.

## Conclusion

Our prospective analyses of the immune status of patients of the ST-ICI cohort demonstrated that LIPS could predict at a very early time point of immune therapy which cancer patients with advanced disease will benefit from ICI treatment and therefore could guide clinical decisions in the future. This newly identified LIPS is a low-cost, safe, easy and broadly applicable and effective early predictor for OS and PFS in metastatic cancer patients treated with ICI.

## Supporting information

sFig1-6

## Data Availability

The datasets used and/or analyzed during the current study are available from the corresponding author on reasonable request.

## Abbreviations

AUC: The area under the ROC curve
CI: Confidence interval
C-index: Concordance index
CTLA-4: Cytotoxic T lymphocyte antigen-4
ECOG: Eastern Cooperative Oncology Group
HNSCC: Head and neck squamous cell carcinoma
HR: Hazard ratios
ICI: Immune checkpoint inhibitors
IPT: immunophenotyping
LASSO: Least absolute shrinkage and selection operator
LIPS: liquid immune profile-based signature
NE: Not estimate
NK cells: Natural killer cells
NKT cells: Natural killer T cells
NSCLC: Non-small cell lung cancer
OS: Overall survival
PBMCs: Peripheral blood mononuclear cells
PD-1: Programmed cell death protein 1
pDCs: Plasmacytoid Dendritic Cells
PD-L1: Programmed cell death ligand 1
PFS: Progression free survival
RECIST: Response Evaluation Criteria in Solid Tumors
ROC: Receiver operating characteristic
TMB: Tumor mutational burden
Tregs: Regulatory T Cells

## Acknowledgements

The authors thank the patients who participated in the study, their supporters, the investigators, and clinical research staff from the Department of Radiation Oncology. The present work was performed by Jian-Guo Zhou in (partial) fulfilment of the requirements for containing the degree “Dr. rer. biol. hum”.

## Contributions

*Conception and design:* MH, USG, BF, RF, HM, JGZ, ME, AD

*Development of methodology:* JGZ, AD

*Acquisition of data:* AD, SR, IB, BF, ME, MH, USG

*Analysis and interpretation of data:* JGZ, AD, ED, RS, PS, USG, MH

*Writing, review, and/or revision of the manuscript:* JGZ, AD, RF, MH, USG, ED, RS, CS, PS

*Administrative, technical, or material support:* SR, CS, MH

*Study supervision:* USG, MH, RF, BF

## Ethics approval and consent to participate

The protocol, any amendments, and informed consent forms were approved by the institutional review boards/independent ethics committees (number: 2_17 B). The study was performed in accordance with the Declaration of Helsinki. All patients gave written informed consent before enrolment that comprised a data privacy clause for data collection and analysis for research purpose.

## Supplementary information

**sFigure 1. Schematic overview of the immune cell types identified in the immunophenotyping assay.**

The scheme depicts the immune cell subtypes analyzed from whole blood. The main cell types (big cells) are identified by their expression of specific cell surface molecules (pan markers). The defined cell types are excluded from the further gating steps, which allows the identification of cell types not expressing characteristic pan markers (dendritic cells, basophils). The main cell types are further divided in cell subtypes (small cells) by analyzing further surface molecules. The immune cell types and subtypes are finally analyzed for the expression of various activation markers (CD25, CD69, CD80, CD86, PD-L1, HLA-DR, CTLA-4 and PD-1).

**sFigure 2. LIPS predict survival benefit from ICI by tumour entity.**

**(A)** The overall survival in HNSCC patients stratified by the LIPS into high- and low-risk with the *P*-value. **(B)** The overall survival in NSCLC patients stratified by the LIPS into high- and low-risk with the *P*-value. **(C)** The progression-free survival in HNSCC patients stratified by the LIPS into high- and low-risk with the *P*-value. **(D)** The progression-free survival in NSCLC patients stratified by the LIPS into high- and low-risk with the *P*-value.

**sFigure 3. LIPS predict survival benefit from ICI by PD-L1 expression.**

**(A)** The overall survival in patients with PD-L1 <1% stratified by the LIPS into high- and low-risk with the *P*-value. **(B)** The overall survival in patients with 1% <PD-L1 < 49% stratified by the LIPS into high- and low-risk with the *P*-value. **(C)** The overall survival in patients with PD-L1 >50% stratified by the LIPS into high- and low-risk with the *P*-value. **(D)** The progression-free survival in patients with PD-L1 <1% stratified by the LIPS into high- and low-risk with the *P*-value. **(E)** The progression-free survival in patients with 1% <PD-L1 < 49% stratified by the LIPS into high- and low-risk with the *P*-value. **(F)** The progression-free survival in patients with PD-L1 >50% stratified by the LIPS into high- and low-risk with the *P*-value.

**sFigure 4. LIPS predict survival benefit from ICI by non-brain metastasis.**

**(A)** The overall survival in patients with brain metastasis stratified by the LIPS into high- and low-risk with the *P*-value. **(B)** The overall survival in patients without brain metastasis stratified by the LIPS into high- and low-risk with the *P*-value. **(C)** The progression-free survival in patients brain metastasis stratified by the LIPS into high- and low-risk with the *P*-value. **(D)** The progression-free survival in patients without brain metastasis stratified by the LIPS into high- and low-risk with the *P*-value.

**sFigure 5. LIPS predict survival benefit from ICI by age.**

**(A)** The overall survival in age < 60yr patients stratified by the LIPS into high- and low-risk with the *P*-value. **(B)** The overall survival in age ≥ 60yr patients stratified by the LIPS into high- and low-risk with the *P*-value. **(C)** The progression-free survival in age < 60yr patients stratified by the LIPS into high- and low-risk with the *P*-value. **(D)** The progression-free survival in age ≥ 60yr patients stratified by the LIPS into high- and low-risk with the *P*-value.

**sFigure 6. LIPS predict survival benefit from ICI by gender.**

**(A)** The overall survival in male patients stratified by the LIPS into high- and low-risk with the *P*-value. **(B)** The overall survival in female patients stratified by the LIPS into high- and low-risk with the *P*-value. **(C)** The progression-free survival in male patients stratified by the LIPS into high- and low-risk with the *P*-value. **(D)** The progression-free survival in female patients stratified by the LIPS into high- and low-risk with the *P*-value.

