## Supplementary figures and images for "Prospective development and validation of a liquid immune profile-based signature (LIPS) to predict response of metastatic cancer patients to immune checkpoint inhibitors"

### Zhou Donaubauer et al_Suppl_Figure 1.pdf

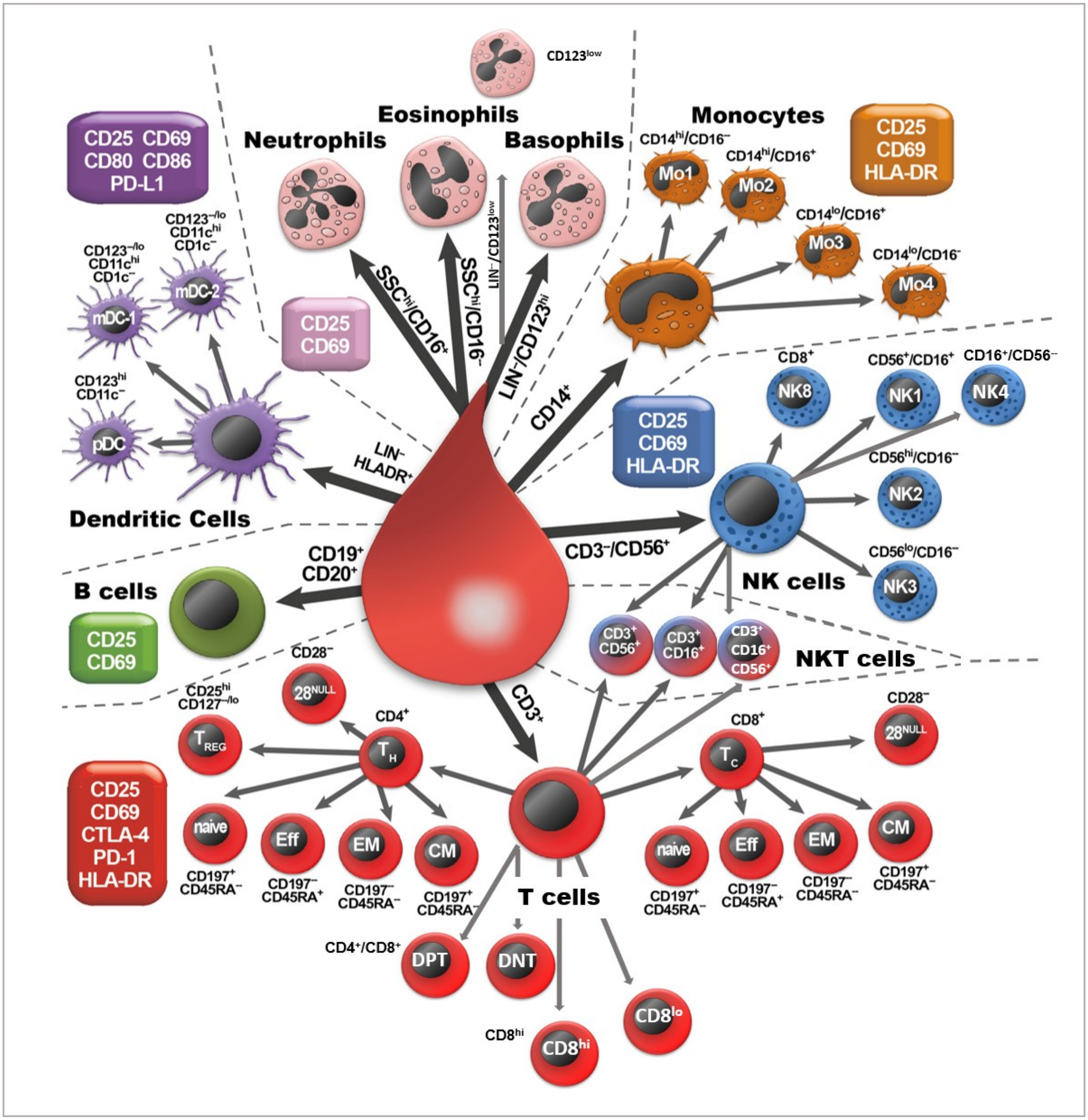

### Zhou Donaubauer et al_Suppl_Figure 2.pdf

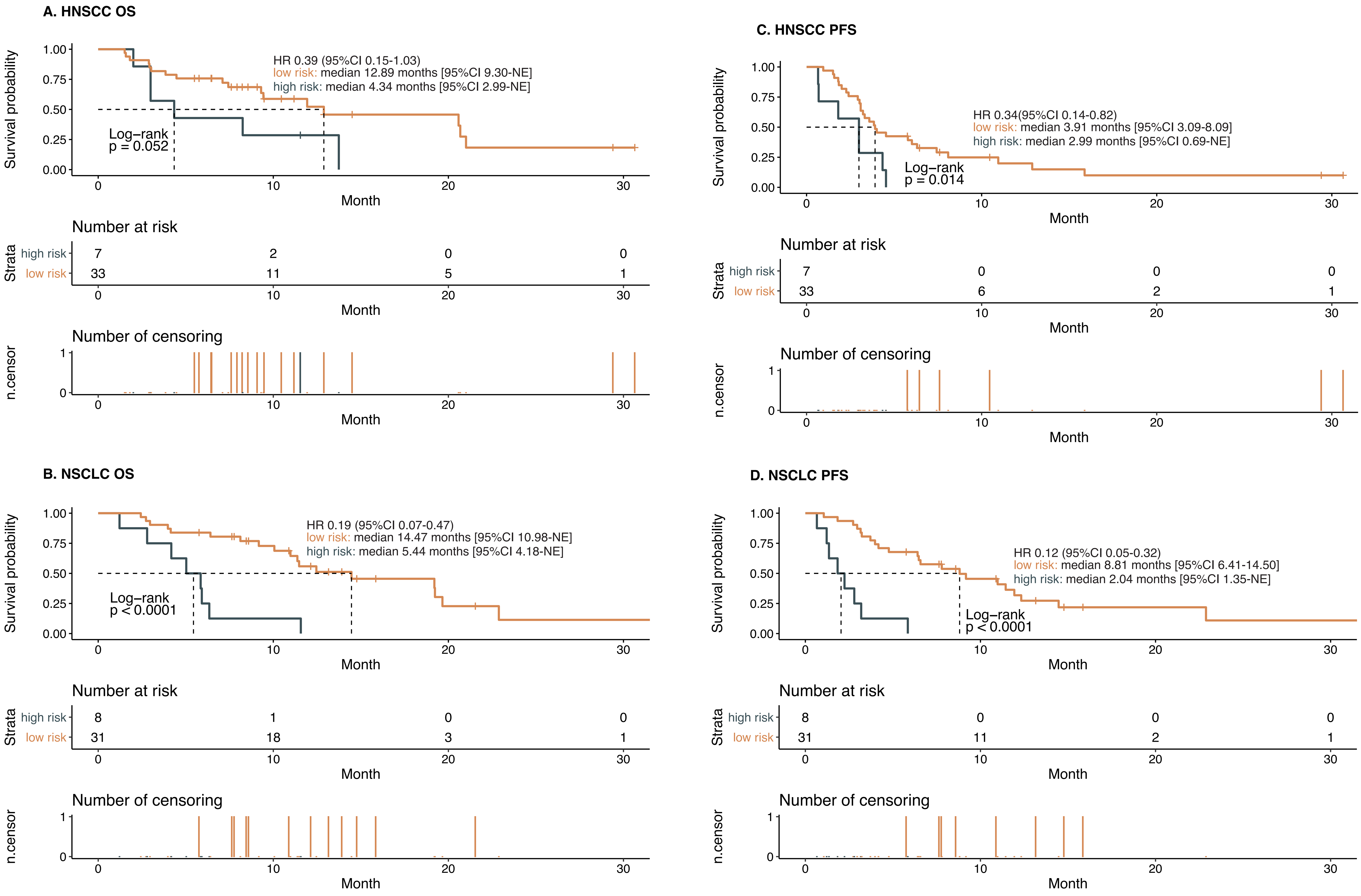

### Zhou Donaubauer et al_Suppl_Figure 3.pdf

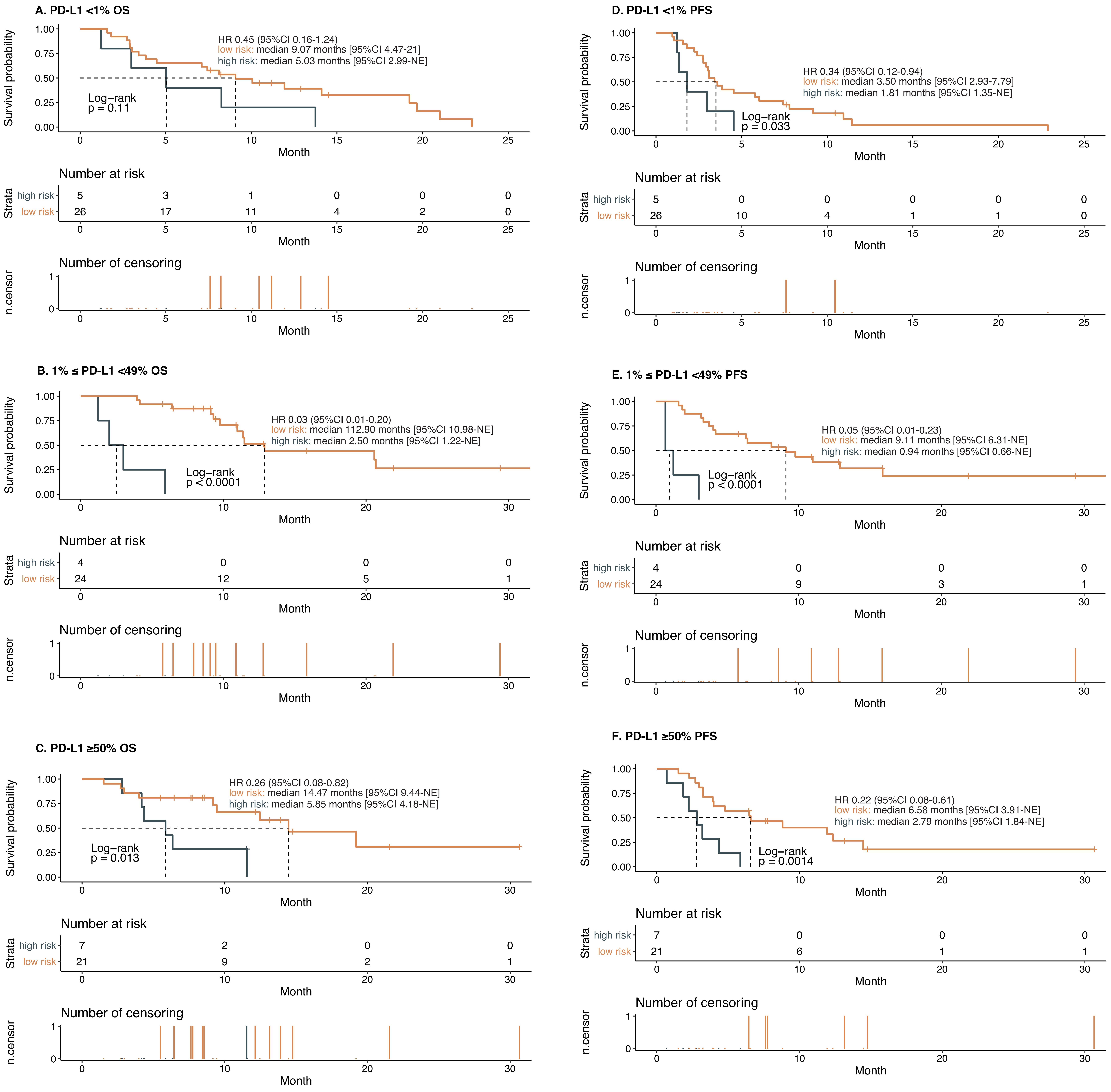

### Zhou Donaubauer et al_Suppl_Figure 4.pdf

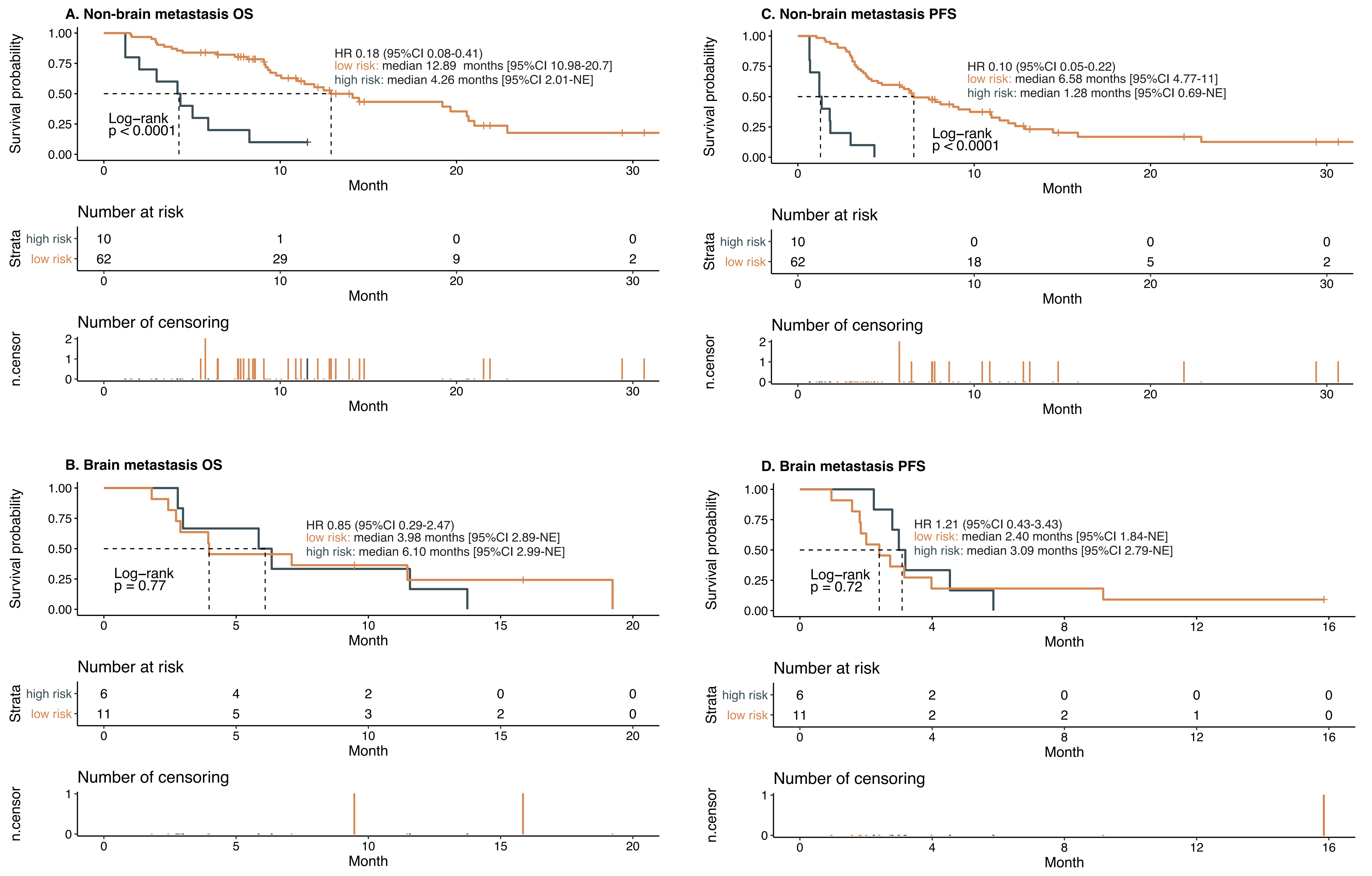

### Zhou Donaubauer et al_Suppl_Figure 5.pdf

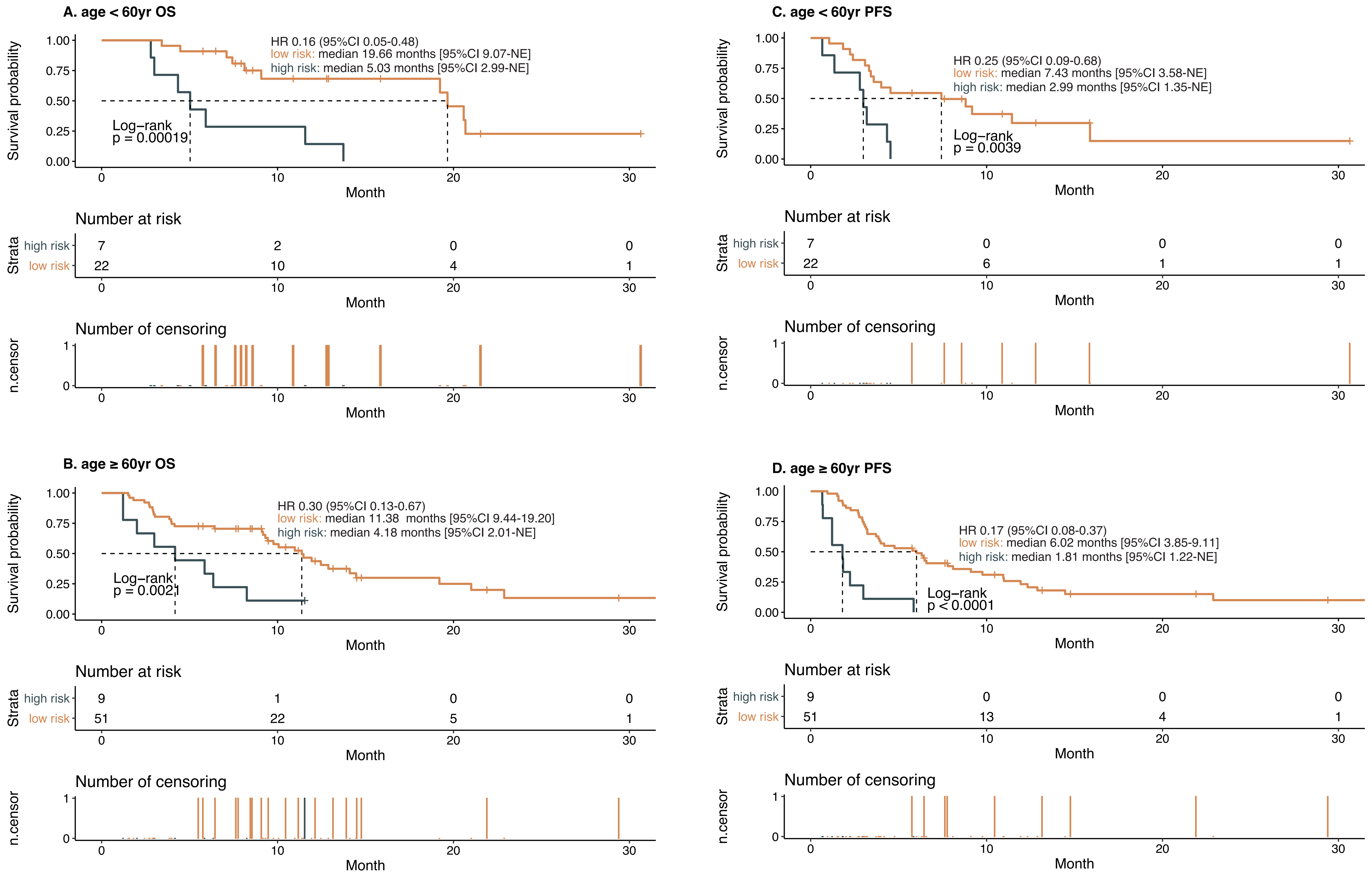

### Zhou Donaubauer et al_Suppl_Figure 6.pdf

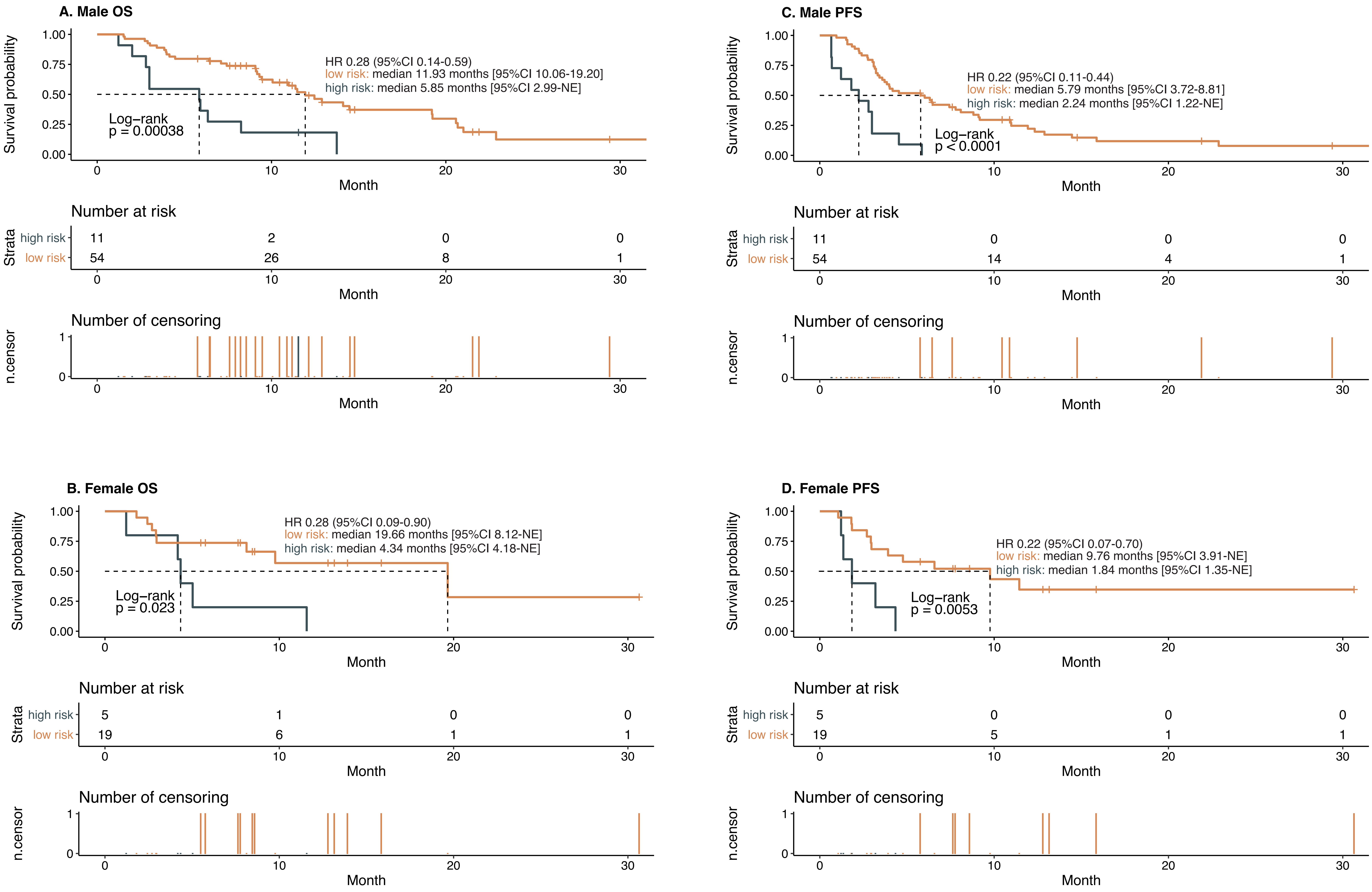
